# ReMIND: The Brain Resection Multimodal Imaging Database

**DOI:** 10.1101/2023.09.14.23295596

**Authors:** Parikshit Juvekar, Reuben Dorent, Fryderyk Kögl, Erickson Torio, Colton Barr, Laura Rigolo, Colin Galvin, Nick Jowkar, Anees Kazi, Nazim Haouchine, Harneet Cheema, Nassir Navab, Steve Pieper, William M. Wells, Wenya Linda Bi, Alexandra Golby, Sarah Frisken, Tina Kapur

## Abstract

The standard of care for brain tumors is maximal safe surgical resection. Neuronavigation augments the surgeon’s ability to achieve this but loses validity as surgery progresses due to brain shift. Moreover, gliomas are often indistinguishable from surrounding healthy brain tissue. Intraoperative magnetic resonance imaging (iMRI) and ultrasound (iUS) help visualize the tumor and brain shift. iUS is faster and easier to incorporate into surgical workflows but offers a lower contrast between tumorous and healthy tissues than iMRI. With the success of data-hungry Artificial Intelligence algorithms in medical image analysis, the benefits of sharing well-curated data cannot be overstated. To this end, we provide the largest publicly available MRI and iUS database of surgically treated brain tumors, including gliomas (n=92), metastases (n=11), and others (n=11). This collection contains 369 preoperative MRI series, 320 3D iUS series, 301 iMRI series, and 356 segmentations collected from 114 consecutive patients at a single institution. This database is expected to help brain shift and image analysis research and neurosurgical training in interpreting iUS and iMRI.

## Background & Summary

Image guidance with computerized navigation based on preoperative magnetic resonance imaging (MRI) was introduced as a surgical adjunct in the 1990s. It facilitates greater resection accuracy by helping the surgeon plan the approach and locate the boundaries of the intended resection. Neuronavigation loses validity as the surgery progresses due to non-linear deformations of the resection cavity and brain shift^1–5^. Intraoperative imaging such as intraoperative MRI (iMRI) and intraoperative ultrasound (iUS) serve to alleviate this issue. While iMRI provides high-contrast images, they can take several minutes up to an hour to acquire. On the other hand, iUS provides relatively lower-contrast images, but they can be acquired within minutes.

Research in the field of medical imaging and image-guided therapy is increasingly seeking large datasets to develop and test machine learning-based algorithms. Specifically, these algorithms aim at improving precision in image guided neurosurgery and interpretability by performing tasks such as image segmentation^6,7^ (e.g., tumor, ventricles, cerebrum, resection cavity), image registration^8^ (e.g., MRI-iMRI, MRI-iUS, iUS-iUS), image synthesis^9^ (e.g., MR to iUS and iUS to iMRI), and visualization^10^ (e.g., uncertainty of registration). However, sharing patient datasets for public research use is challenging due to the significant resources required for data curation as well as the need to ensure patient privacy. Moreover, datasets that combine multimodal imaging from both preoperative and intraoperative acquisitions in the same patient are particularly scarce. To address this gap, we have curated a database from neurosurgical procedures conducted in the Advanced Multimodality Image Guided Operating (AMIGO) suite at the Brigham and Women’s Hospital. This database represents the largest publicly-available collection of preoperative MRI, intraoperative MRI, and intraoperative ultrasound data from surgically treated brain tumors. It contains 92 gliomas, 11 metastases, and 11 non-glioma pathologies. The database includes 369 preoperative MRI series, 320 three-dimensional iUS sweeps, 301 iMRI series, and 356 segmentations obtained from 114 consecutive patients who underwent image-guided resection at a single institution. Additionally, each case contains segmentations of the preoperative tumor, the pre-resection cerebrum, and the previous resection cavity derived from the preoperative MRI (if applicable), as well as any residual tumor identified on the iMRI. For reference, Figure 1 provides an illustrative example of the contents of each dataset and all available segmentations.

**Figure 1.**
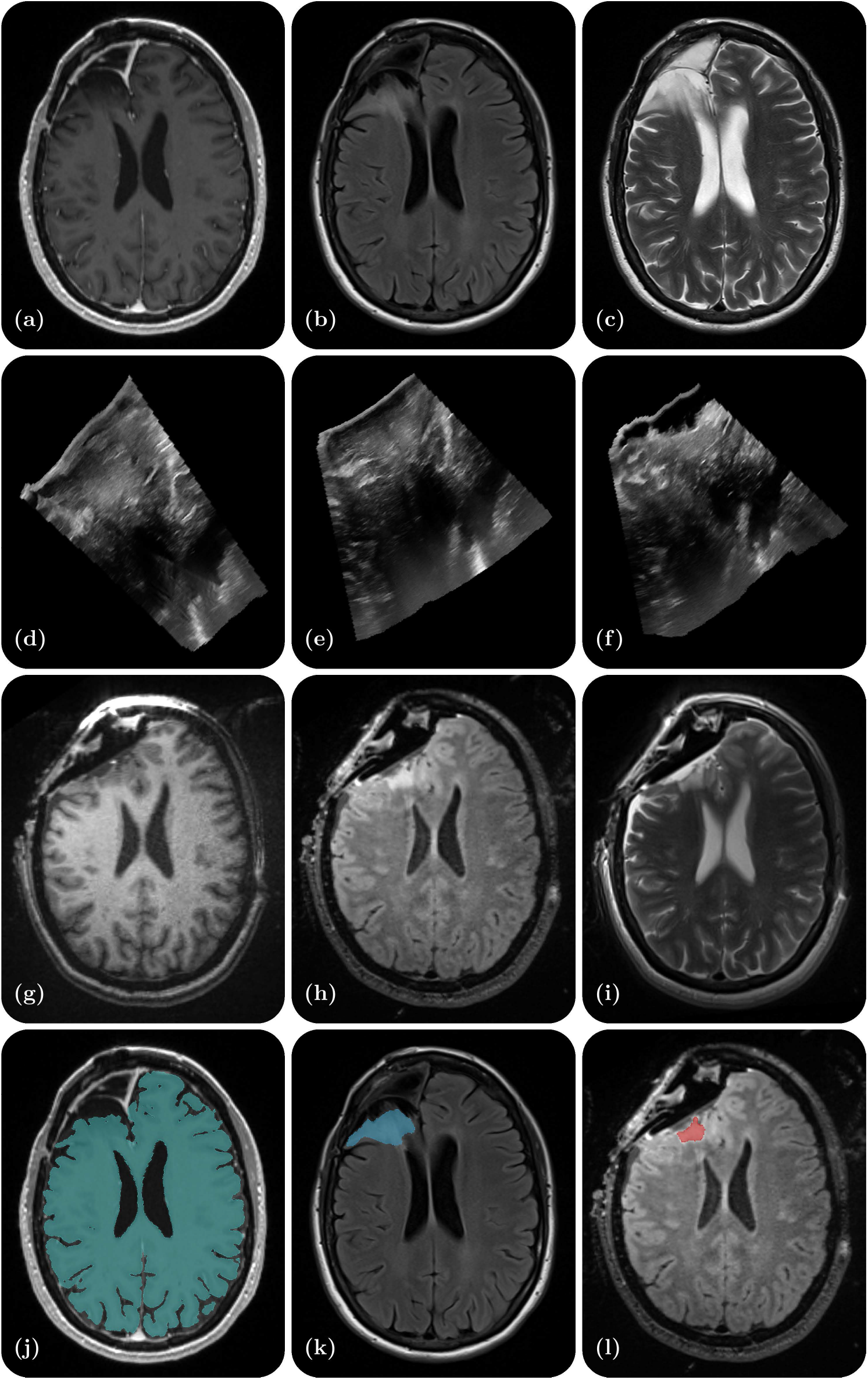
Illustrative example of one dataset - a right frontal lobe recurrent WHO Grade II Oligodendroglioma (IDH-positive, 1p/19q co-deleted). (a) Preoperative contrast-enhanced T1-weighted MR; (b) Preoperative T2-weighted MR; (c) Preoperative T2 FLAIR MR; (d) Intraoperative US prior to dural opening; (e) Intraoperative US post dural opening; (f) Intraoperative US prior to iMRI; (g) Intraoperative contrast-enhanced T1-weighted MRI; (h) Intraoperative T2 FLAIR MRI; (i) Intraoperative T2-weighted MRI (BLADE); (j) Cerebrum segmentation on the preoperative contrast-enhanced T1-weighted MRI; (k) Tumor segmentation on the preoperative T2 FLAIR MRI; (l) Residual tumor segmentation on the intraoperative T2 FLAIR MRI.

With this work, we build upon the effort initiated by the Montreal Neurosurgical Institute (MNI) and St. Olav University Hospital (Trondheim, Norway) to publicly share MRI and iUS images acquired for brain tumor patients through the BITE^11–13^ (Brain Images of Tumors for Evaluation) and RESECT^14,15^ (REtroSpective Evaluation of Cerebral Tumors) databases (Table 1) respectively.

**Table 1.** Comparison against existing publicly available databases of preoperative and intraoperative imaging in brain cancer patients.

|  | MNI BITE<sup>11-13</sup> | RESECT<sup>14,15</sup> | ReMIND |
| --- | --- | --- | --- |
| Group | Montreal | Trondheim | Boston |
| Publication Year(s) | 2012<sup>11</sup>, 2014<sup>12</sup>, 2016<sup>13</sup> | 2017<sup>14</sup>, 2022<sup>15</sup> | 2023 |
| Patients | 14<sup>12</sup> (+9<sup>13</sup>) | 23 | 114 |
| Tumor Type(s) | Low-Grade Gliomas<br>High-Grade Gliomas | Low-Grade Gliomas | Low-Grade Gliomas<br>High-Grade Gliomas<br>Other tumors |
| Preoperative MRI | 1.5T GE Signa EXCITE<br>3T Siemens Magnetom TIM Trio | 1.5 Siemens Magnetom Avanto<br>3T Siemens Magnetom Skyra | Various 1.5T/3T/7T scanners<br>(Siemens, GE, Hitachi) |
| Intraoperative MRI (iMRI) | - | - | 3T Siemens Magnetom Verio<br>70 cm Wide-Bore |
| Postoperative MRI | 1.5T GE Signa EXCITE | - | - |
| Smallest MRI Voxel Size | 1 × 0.5 × 0.5 mm | 1 × 1 × 1 mm | 1 × 1 × 0.5 mm |
| Neuronavigation<br>Processing Unit | IBIS NeuroNav | Sonowand Invite | Brainlab Curve Dual Display<br>Brainlab Cranial<br>Navigation, Brainlab Ultrasound |
| Neuronavigation Optical<br>Tracking Unit | Polaris<br>(Northern Digital Inc.) | Polaris Spectra<br>(Northern Digital Inc.) | Polaris Spectra for Brainlab<br>(Northern Digital Inc.) |
| Navigation Tracking<br>Adapter for iUS Probe | TA003<br>(Traxtal Technologies Inc.) | Built-in | Adapter Base (41860-XX)<br>Array (22595) (Brainlab AG) |
| iUS Processing Unit | HDI 5000<br>(Philips ATL) | Built-in | bk5000<br>(BK medical) |
| iUS Probe | 4-7 Mhz Phased<br>Array Transducer P7-4<br>(Philips ATL) | 12-6 Mhz Flat Linear<br>Array Transducer 12FLA-L<br>(Sonowand AS) | 13-5 Mhz Curved Array<br>Array Transducer N13C5<br>(9062) (BK medical) |
| iUS Image Depth | 6.5 cm or 8 cm | - | 6.5 cm |
| iUS Reconstruction<br>Resolution | 2D: 0.2 × 0.2 mm<br>3D: 0.3 × 0.3 × 0.3 mm | 0.14 × 0.14 × 0.14 mm to<br>0.24 × 0.24 × 0.24 mm | 0.1 × 0.1 × 0.5 mm |
| Extradural iUS<br>(before dural incision) | 2D & 3D | 3D | 3D |
| Intradural iUS<br>(after dural incision) | - | - | 3D |
| Resection Control iUS | - | 3D | - |
| Post-resection (pre-iMRI) iUS | 2D & 3D | 3D | 3D |
| Segmentations | Preoperative Tumor (Manual) | Preoperative Tumor (Manual)<sup>15</sup><br>Resection cavity in iUS (Manual) | Preoperative Tumor (Manual)<br>Prior Resection Cavity (Manual)<br>Preoperative Ventricles (Auto)<br>Preoperative Cerebrum (Auto) |
| Landmarks | iUS to iUS<br>post-resection iUS to MRI | iUS to iUS<br>pre-resection iUS to MRI<sup>14</sup> | Not yet available |

## Methods

This section describes all the procedures followed to acquire and curate the data within the Brain Resection Multimodal Imaging Database (ReMIND) collection^16^, including experimental design, data acquisition, data annotation, and computational processing (e.g., format conversion, defacing).

### Patient cohort

The ReMIND database^16^ comprises 123 consecutive patients who were surgically treated with image-guided tumor resection in the AMIGO Suite at the Brigham and Women’s Hospital (Boston, USA), between November 2018 and August 2022, using both intraoperative ultrasound (iUS) and intraoperative MRI (iMRI). Of the 123 cases, 9 were excluded due to corrupted or poor-quality data, resulting in a final cohort of 114 cases. Specifically, the reasons for exclusion include the presence of large artifacts in iMRI (N=1), the corruption of 3D iMRI (N=1), or poor quality of intraoperative US prior to dural opening (N=7). Summary demographics can be found in Table 2.

**Table 2.** Summary of the clinical metadata of the ReMIND data collection.

| Clinical Data (N=114) |  |  |  |
| --- | --- | --- | --- |
| **Demographics** |  | **Histopathology** |  |
| Age | 47.50 [32.75 – 59.00] | Gliomas, glioneuronal, and neuronal tumors | 83.3% (95/114) |
| Sex |  | *Astrocytoma* | 34.7% (33/95) |
| *Male* | 53.5% (61/114) | *Glioblastoma* | 32.6% (31/95) |
| *Female* | 46.5% (53/114) | *Oligodendroglioma* | 25.3% (24/95) |
| Race |  | *Other* | 7.4% (7/95) |
| *White* | 90.4% (103/114) | Meningiomas | 0.9% (1/114) |
| *Asian* | 5.3% (6/114) | Hematolymphoid tumors | 0.9% (1/114) |
| *Declined* | 2.6% (3/114) | Metastases | 9.6% (11/114) |
| *Other* | 1.8% (2/114) | Other | 5.3% (6/114) |
| Ethnicity |  | **Adult-type diffuse gliomas (n=88)** |  |
| *Not Hispanic* | 94.7% (108/114) | **WHO grade** |  |
| *Hispanic* | 4.4% (5/114) | 1 | 3.2% (3/88) |
| *Declined* | 0.9% (1/114) | 2 | 31.6% (31/88) |
| **Preoperative MRI** |  | 3 | 24.2% (23/88) |
| Tesla |  | 4 | 38.9% (37/88) |
| 3T | 71.1% (81/114) | Not assigned | 2.1% (2/88) |
| 1.5T | 21.1% (24/114) | **Molecular genetics** |  |
| 7T | 1.8% (2/114) | IDH (n=88) |  |
| 1.16T | 1.8% (2/114) | *IDH-mutant* | 64.8% (57/88) |
| Eloquent |  | *IDH-wildtype* | 35.8% (31/88) |
| *Yes* | 67.5% (77/114) | 1p/19q (n=75) |  |
| *No* | 32.5% (37/114) | *retained* | 65.3% (49/75) |
| Enhancement | 57.9% (66/114) | *co-deleted* | 33.3% (25/75) |
| T2/FLAIR hyperintensity | 64.0% (73/114) | *1p-retained, 19q-deleted* | 1.3% (1/75) |
| Volume (cm3) | 17.83 [6.31 – 35.87] | MGMT promoter (n=83) | 94.3% (83/88) |
| Tumor depth |  | *Methylated* | 56.6 % (47/83) |
| *Gyral and subgyral* | 49.1% (56/114) | *Unmethylated* | 34.9% (29/83) |
| *Subgyral* | 33.3% (38/114) | *Partially methylated* | 8.4% (7/83) |
| *Gyral* | 17.5% (20/114) |  |  |
| **Surgical parameters** |  |  |  |
| Previous craniotomy |  |  |  |
| *No* | 55.3% (63/114) |  |  |
| *Yes* | 44.7% (51/114) |  |  |
| Anesthesia technique |  |  |  |
| *General anesthesia* | 95.6% (109/114) |  |  |
| *Awake craniotomy* | 4.4% (5/114) |  |  |
| Laterality |  |  |  |
| *Left* | 50.0% (57/114) |  |  |
| *Right* | 50.0% (57/114) |  |  |
| Resection after iMRI | 71.1% (81/114) |  |  |
| Complete ultrasounds | 80.7% (92/114) |  |  |

The patients in the study were treated according to the current standard of care, augmented with intra-operative imaging. Collection, analysis, and release of the ReMIND database have been performed in compliance with all relevant ethical regulations. The Institutional Review Board at the Brigham and Women’s Hospital approved the protocol (2002-P-001238), and informed consent was obtained from all participants, including for public sharing of data.

### Surgical Setup with Neuronavigation

Neuronavigation systems aim at providing intraoperative guidance to neurosurgeons by allowing them to visualize the position of their surgical tools relative to the tumor and critical brain areas visible in preoperative Magnetic Resonance Imaging (MRI). Specifically, intraoperative neuronavigation was performed using the optical tracking version of the “Curve” Dual Display system (Brainlab AG, Munich, Germany). The patient reference frame was established by rigidly securing the MRI-safe “Standard Cranial Reference Array with 4 Marker Spheres” (Brainlab AG, Munich, Germany) to the MRI-safe IMRIS head holder (IMRIS Inc., Minnesota, USA). Patients were positioned either supine or lateral, with variable head positions ranging from neutral to approximately 90 degrees to one side. No patients were positioned prone.

The image-to-patient registration was performed in two steps. In the first step, the precalibrated “Softouch Pointer” (Brainlab AG, Munich, Germany) was used to perform an initial image-to-patient registration with the “Cranial Navigation” module (Brainlab AG, Munich, Germany). The nasion, left lateral canthus, and right lateral canthus were located on the patient using the tracked pointer to establish an initial registration. In the second step, a dense set of points was acquired on the skin to refine the patient registration to the skin surface on preoperative imaging. For this step, the “Cranial Navigation” employs a registration approach based on the iterative closest point algorithm with a custom point-to-surface distance function^17^. Surface-based registration was consistently achieved in all cases by ensuring that the patient’s face was visible to the camera during registration. Finally, the quality of the registration was visually verified by the surgeon and improved when necessary by collecting additional registration points.

### Clinical, Demographic, and Pathology data

Demographic information, including age, sex, and ethnicity, was obtained from the corresponding patient medical records. The age range of the included population was 20–76. The ratio of male:female was equal to 61:53. Moreover, clinico-pathologic data such as the tumor type, tumor grade, radiological characteristics upon contrast administration, tumor location, and the reoperation status were assessed by the treating neurosurgeons. Tumor type and grade were specified according to the World Health Organization (WHO) 2021 Classification of Tumors of the Central Nervous System.^18–21^. Additionally, tumors were classified into one of 3 categories based on proximity to the functional cortex (non-eloquent, near eloquent, and eloquent).

Table 2 shows a summary of the clinical metadata. A number of the surgeries were reoperations (44.7%) and a minority were performed awake (4.4%). There was an equal number of right- and left-sided craniotomies. The majority of the patients treated were classified as gliomas, glioneuronal, and neuronal tumors (83.3%) and were primarily Astrocytomas (34.7%). IDH-mutations, 1p/19q-codeletions, and MGMT promoter methylation status were reported in 100%, 67.5%, and 94.3% of the adult-type diffuse gliomas, respectively. The majority of the gliomas, glioneuronal, and neuronal tumors were CNS WHO Grade 4 (38.9%). (67.5%) were located in eloquent areas based on the fMRI, anatomic substrate, and neurophysiological monitoring. The full metadata table can be downloaded from our repository^16^ on The Cancer Imaging Archive (TCIA).

### Imaging data

The ReMIND database^16^ corresponds to a collection of preoperative MRI, intraoperative MRI, and intraoperative ultrasounds. A summary of the acquisition parameters for preoperative and intraoperative MRI is provided in Table 1. The imaging data can be found and downloaded from our TCIA repository^16^ as well.

#### Preoperative MRI

Preoperative MRI were accessed, selected, and co-registered using the “Elements” software (Brainlab AG, Munich, Germany), a CE-certified / FDA-cleared medical device software used routinely in clinical practice. Specifically, “Image Fusion”, a mutual information-based registration algorithm^22^, was used to rigidly co-register preoperative images. Preoperative MRI comprises four structural MRI sequences: native T1-weighted (T1), contrast-enhanced T1-weighted (ceT1), native T2-weighted (T2), and T2-weighted fluid-attenuated inversion recovery (T2-FLAIR). These scans were acquired before surgery using various scanners at multiple institutions, making their acquisition parameters heterogeneous, as shown in Table 1. More details about the acquired sequences are presented in Table 3. Most of the preoperative imaging was performed on a 3T (71.1%) MRI scanner with Siemens (87.7%) being the most common manufacturer.

**Table 3.** Summary of data characteristics of the ReMIND MR sets.

| Pre-operative MR |  |  |  |  |  |  |  |
| --- | --- | --- | --- | --- | --- | --- | --- |
|  | MP-RAGE T<sub>1</sub> | MP2RAGE T<sub>1</sub> | MP-RAGE ceT<sub>1</sub> | SPACE T<sub>2</sub> | BLADE T<sub>2</sub> | FLAIR T<sub>2</sub> | Other T<sub>2</sub> |
| Acquisition | 3D | 3D | 3D | 3D | 2D | 2D (90%)<br>3D (10%) | 2D (81%)<br>3D (19%) |
| Patient availability | 5 (4%) | 9 (8%) | 105 (92%) | 71 (62%) | 34 (30%) | 40 (35%) | 82 (72%) |
| Average slice number | 191 ± 10 | 175 ± 9 | 194 ± 38 | 186 ± 12 | 72 ± 23 | 53 ± 63 | 66 ± 66 |
| In-plane resolution in mm | 1.0 ± 0.0 | 1.0 ± 0.0 | 0.9 ± 0.2 | 0.9 ± 0.1 | 0.6 ± 0.2 | 0.4 ± 0.2 | 0.7 ± 0.2 |
| Slice thickness in mm | 1.0 ± 0.0 | 1.0 ± 0.0 | 1.0 ± 0.1 | 1.0 ± 0.1 | 2.5 ± 0.8 | 4.5 ± 1.5 | 4.0 ± 1.7 |

| Intra-operative MR |  |  |  |  |  |  |  |
| --- | --- | --- | --- | --- | --- | --- | --- |
|  | MP-RAGE T<sub>1</sub> | MP2RAGE T<sub>1</sub> | MP-RAGE ceT<sub>1</sub> | SPACE T<sub>2</sub> | BLADE T<sub>2</sub> | FLAIR T<sub>2</sub> | Other T<sub>2</sub> |
| Acquisition | 3D | 3D | 3D | 3D | 2D | 2D | 2D (29%)<br>3D (71%) |
| Patient availability | 19 (17%) | 8 (7%) | 63 (55%) | 5 (4%) | 108 (95%) | 1 (1%) | 77 (68%) |
| Average slice number | 176 ± 0 | 180 ± 7 | 176 ± 5 | 179 ± 6 | 70 ± 2 | 40 ± 0 | 145 ± 47 |
| In-plane resolution in mm | 1.0 ± 0.0 | 1.0 ± 0.0 | 1.0 ± 0.0 | 0.8 ± 0.2 | 0.9 ± 0.0 | 0.7 ± 0.0 | 0.5 ± 0.0 |
| Slice thickness in mm | 1.2 ± 0.1 | 1.0 ± 0.0 | 1.1 ± 0.1 | 1.0 ± 0.0 | 2.1 ± 0.3 | 3.9 ± 0.0 | 1.7 ± 1.0 |

#### Intraoperative MRI (iMRI)

Unlike preoperative MRI, all intraoperative MRI were acquired in the AMIGO suite at the Brigham and Women’s Hospital (Boston, USA) using a 3T wide-bore (70 cm) MRI scanner (Magnetom Verio, Siemens Healthineers, Erlangen, Germany) with an 8-channel head coil to evaluate the presence of residual targeted tissue once the planned portion of targeted tissue was removed. Before the acquisition, a temporary closure of the craniotomy was performed. The entire acquisition process required 1-1.5 hrs. and included MRI safety procedures, instrument counts, preparation for scanning, and redraping to resume surgery after iMRI.

#### Intraoperative Ultrasound (iUS)

All iUS series were acquired using a sterilizable 2D neuro-cranial curvilinear transducer on a cart-based ultrasound system (N13C5, BK5000, GE Healthcare, Peabody, MA, USA) in the AMIGO suite. The ultrasound probe had a contact area of 29mm × 10mm and a frequency range of 5-13 MHz. The “Ultrasound Navigation Adapter Array” together with the “Ultrasound Navigation Adapter Base - BK N13C5” (Brainlab AG, Munich, Germany) were attached to the iUS probe to enable the Curve platform to track the probe relative to the patient. The imaging plane was chosen to be as parallel as possible to one of the three cardinal axes of the head (axial, sagittal, coronal). However, this was often limited by the size and shape of the craniotomy. The transducer was swept unidirectionally at a slow, consistent speed through the craniotomy. This specific motion, in conjunction with the tracking, enabled the reconstruction of a 3D volume from the tracked 2D sweeps using the “Ultrasound” module within the “Elements” software platform on the “Curve” hardware system. Similar to the RESECT database^14^, 3D iUS acquisition was aimed to be performed at three distinct surgical time points:

1. **Pre-dura iUS:** between the craniotomy and the dural opening.
2. **Post-dura iUS:**after the dural opening but before any tumor resection was performed.
3. **Pre-iMRI iUS:** immediately before the iMRI was acquired, i.e. after substantial tumor resection was completed to the degree that either the surgeon was satisfied with the microscopically-visible extent of resection or that iMRI was needed to identify the remaining portion of the tumor.

When more than one iUS volume was acquired at a specific surgical time point, the acquisition with the best image quality, field of view, and maximal tumor coverage was included in this collection. In 22 cases, iUS acquisitions were unavailable at some surgical time points due to surgeon preference (15/22) or durotomies during craniotomies or previous surgeries (7/22). Such cases have an explanation detailed in the clinical metadata table available on TCIA.

### Segmentation data

Various segmentations were created to assist the surgical resection. These typically include manual segmentations of the preoperative whole tumor, preoperative tumor target (i.e., the radiologically identifiable tumor specifically targeted for resection), resection cavity resulting from prior surgery (i.e. in case of reoperation) and intraoperative residual tumor. These segmentations were performed using the BrainLab “Elements” planning software by a neurosurgical fellow and refined if needed by the attending neurosurgeon. For enhancing lesions, the preoperative and residual tumor segmentation encompass the enhancing region and any necrotic core if present. In the case of non-enhancing lesions, the preoperative and residual tumor segmentations correspond to the solid/nodular component within the T2/T2-FLAIR hyperintensity region.

Moreover, automatic segmentations of cerebrum and ventricles were obtained using the “Object Manipulation” module (Brainlab AG, Munich, Germany). The segmentation of these two structures serves both clinical and research purposes. The three-dimensional model of the cerebrum helps neurosurgeons to assess the accuracy of the navigation using gyral anatomy and correct any small errors and to select the operative trajectory. Additionally, the segmentation of ventricles is useful to assess registration and any brain shift since they are readily visible on iUS; clinically these can be helpful for avoiding inadvertent entry during procedures. Only structures deemed necessary for surgical resection by the attending neurosurgeon were automatically segmented. For research purposes, the segmentation of the cerebrum and ventricles may be valuable as the starting point for the development and validation of algorithms, such as those for brain structure segmentation in the presence of lesions or image registration.

In total, manual preoperative whole tumor segmentations are provided for 113 cases; preoperative tumor target segmentations are provided for 3 cases in which subtotal resection was planned; manual previous resection cavity segmentations are provided for 21 cases; residual tumor segmentations are provided for 58 cases; and automated segmentations of the cerebrum and ventricles are provided for 89 and 54 cases respectively. All cerebrum, ventricle, and tumor segmentations were created preoperatively during the surgical planning stage. In contrast, residual tumor segmentations were created intraoperatively from iMRI.

### Data export

Preoperative MRI were accessed, selected, and co-registered using the “Image Fusion” module within the “Elements” software platform (Brainlab AG, Munich, Germany). After visual inspection, some MR series were excluded due to poor quality, the presence of artifacts, or a small field of view. Manual and automated segmentations were performed using the “Object Manipulation” module. Intraoperative ultrasounds were tracked using the “Curve” neuronavigation system, allowing them to be roughly registered with the preoperative images. Finally, intraoperative MRI were automatically registered with preoperative scans also using the “Image Fusion” module in “Elements”. Note that image registration was performed by only updating image headers to avoid resampling errors. Images and segmentations were finally exported as NRRD files from the “Curve” neuronavigation system using 3D Slicer via OpenIGTLink^23^.

### Data transmission, de-identification and format conversion

The de-identified imaging data and segmentation data were submitted to The Cancer Imaging Archive (TCIA) in DICOM format (imaging and segmentation) and NRRD format (segmentation). Experienced quality-control reviewers inspected images to ensure the data are fully de-identified and well-curated.

Data were fully de-identified by removing all health information identifiers and by applying a de-facing algorithm. Specifically, each MR scan was defaced by: 1/ affinely registering an MR template to the MR scan using NiftyReg^24^; 2/ applying the obtained affine transformation to the face mask of the template; 3/ applying the resampled face mask to the MR scan. The code of the algorithm is publicly available at https://github.com/ReubenDo/pydeface-niftyreg. All the defaced scans were visually inspected, and 100% of them were successfully defaced.

The DICOM format is increasingly used in research as it is standardized, preserves all metadata (e.g. patient, session), and allows for long-term archival. For that reason, our imaging data was released in DICOM format on TCIA. Specifically, the preoperative and intraoperative MR images were converted from NRRD to DICOM series using 3D Slicer^25^. Moreover, the reconstructed 3D iUS images were converted to multi-frame DICOM using the dicom3tools software^26^. Finally, each segmentation was converted using DCMqi^27^ with the segmented MR DICOM data as the reference image.

## Data Records

All the imaging data and the metadata described here as the “ReMIND” collection are available as a publicly available repository of The Cancer Imaging Archive (TCIA)^28^ (TCIA: https://doi.org/10.7937/3RAG-D070)^16^. The DICOM files conform to DICOM standards and are organized on TCIA based on anonymized patient ID numbers (CaseID). Each patient is associated with two studies: preoperative and postoperative, each comprising images and segmentations. Note that ultrasound images are exclusively part of the intraoperative study. As a result, the downloaded DICOM database follows the folder structure outlined in Box 1. Segmentation NRRD files are organized similarly, as depicted in Box 2.

### Box 1.

**Folder structure of the DICOM TCIA dataset**

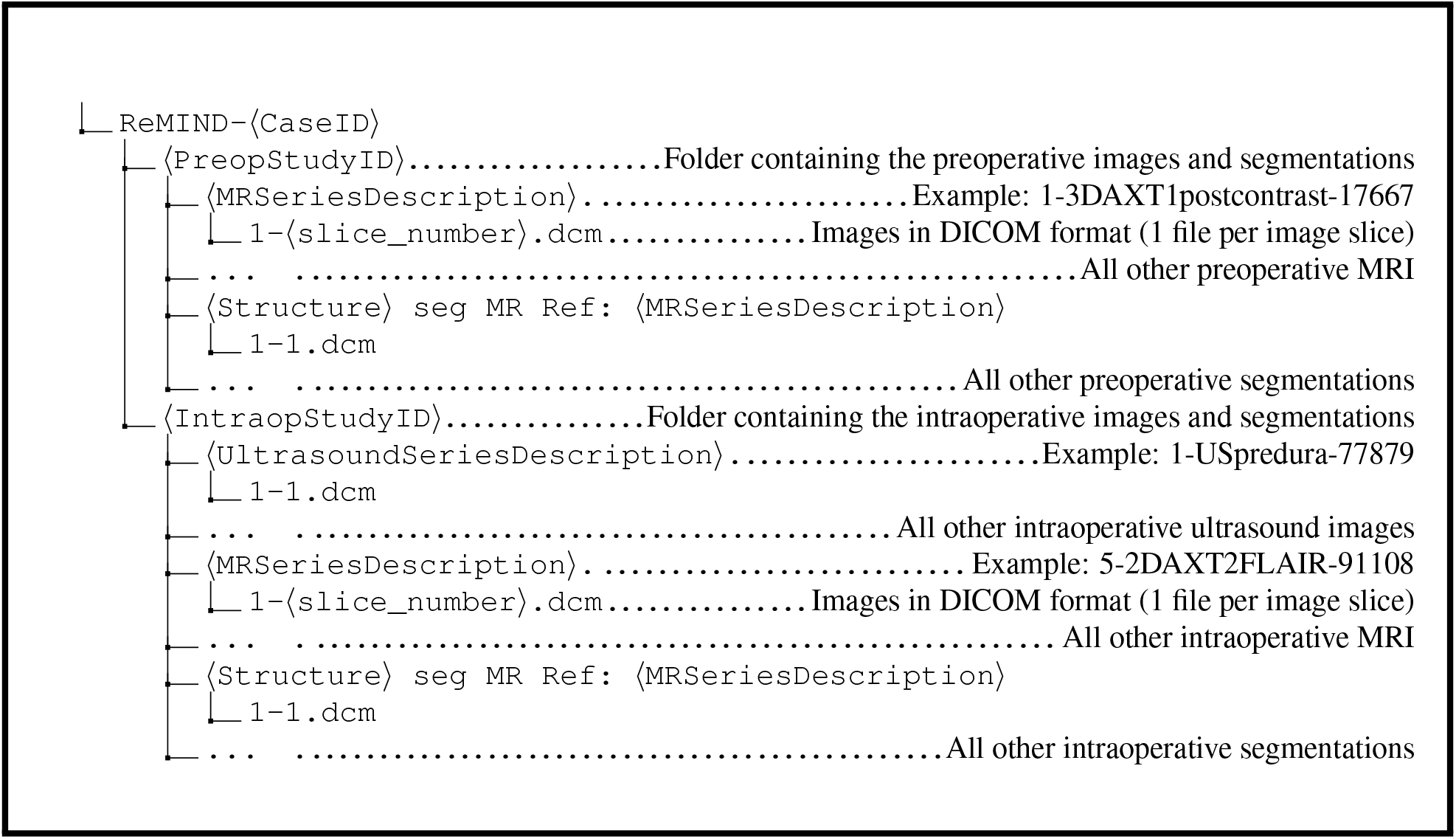

### Box 2.

**Folder structure of the NRRD segmentation dataset.**

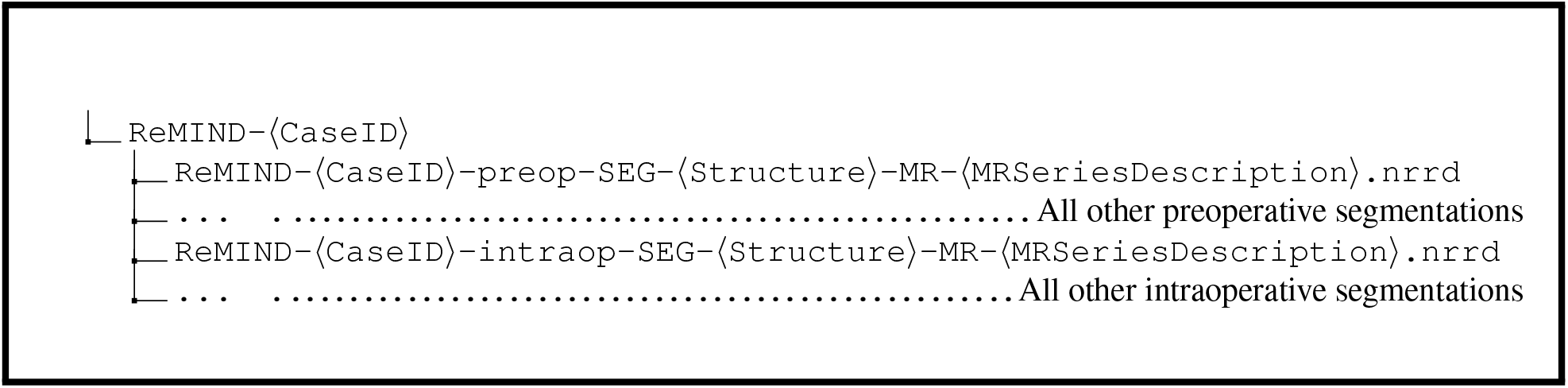

## Technical Validation

Quality control steps are performed before, during, and after surgery. These include assessing image quality as well as quality of image-to-image registration and image-to-patient registration, image deidentification, segmentation, and data conversion. We present these steps below.

### Image selection and registration steps

During surgical planning, a neurosurgical fellow selected the preoperative MR data included in the surgical plan. These images correspond to the ReMIND’s preoperative data. In particular, the fellow ensured data integrity by checking for corruption or significant artifacts. Selected preoperative scans were then co-registered using the Brainlab “Image Fusion” module. The neurosurgical fellows and the attending neurosurgeon visually assessed the registration quality during surgical planning, confirming successful co-registration for all cases.

### Image-to-Patient Registration

At the start of the procedure, image-to-patient registration was performed and assessed by the attending neurosurgeon using surface landmarks. If needed, the neurosurgeon collected additional registration points on the patient’s skin surface to refine the registration. The surgeons assessed the registration accuracy by visually checking the position of the tracked pointer with surface landmarks on the pre-operative MRI. If a substantial inaccuracy was observed, the registration step was repeated until acceptable accuracy was obtained.

### Image de-facing

The derived de-facing masks for all cases were visually checked for quality by a research fellow and experienced quality-control TCIA reviewers. The developed tool was robust with 100% of the preoperative and intraoperative MR scans successfully defaced.

### Imaging data conversion

To ensure that the data conversion step (from NRRD to DICOM) did not deteriorate raw imaging data, DICOM images were converted back to NRRD files using 3D Slicer. Then, the 3D image arrays of the original NRRD and the converted NRRD files were compared using the Simple Insight Toolkit^29^ (SimpleITK). The conversion process was found to preserve data information, i.e. the same values were found at each voxel location.

### Automated Segmentation Data

Automated brain structure segmentations were subject to visual inspection around the surgical areas of interest by the neurosurgical fellows and the attending neurosurgeon during surgical planning. Although these automated segmentations were generated by a widely trusted commercial software routinely used in clinical practice, inaccuracies may occur, especially for cases involving reoperations of recurrent tumors. For that reason, these segmentations should not be used as a gold-standard for segmentation problems, but would serve well as starting points which could be further refined.

### Manual Segmentation Data

Preoperatively, the tumor annotations were performed by the neurosurgical fellow during surgical planning. These annotations were assessed by the attending neurosurgeon and refined if needed. Intraoperatively, the neurosurgical fellow and the attending neurosurgeon segmented the residual tumor visible in intraoperative MRI. As part of the public release of this collection, interobserver variability was measured for the manual tumor segmentations on 10 cases (5 enhancing and 5 non-enhancing tumors) for further validation. Preoperative T1 gadolinium enhanced MRI was used to segment enhancing tumors and preoperative T2-weighted MRI (SPACE) were used for non-enhancing tumors were segmented independently by two neurosurgical research fellows. We quantitatively measured the Dice score and average symmetric surface distance (ASSD) between the two sets of manual segmentations. For manual tumor segmentation on enhanced T1-weighted MRI, interobserver variability yielded a Dice score of 92.2% (SD 2.8%) and an ASSD score of 0.68mm (SD 0.34mm). For manual non-enhancing tumor segmentation on T2-weighted MRI, interobserver variability yielded a Dice score of 86.4% (SD 3.7%) and an ASSD score of 1.24mm (SD 0.22mm).

## Data Availability

All data produced are available online at https://doi.org/10.7937/3rag-d070

## Usage Notes

To view the imaging data, we recommend 3D Slicer available at https://www.slicer.org, which is a free and open-source platform for medical image informatics, image processing, and three-dimensional visualization. 3D Slicer supports the DICOM format. We also developed a custom 3D Slicer extension to create landmarks on images. The MRUSLandmarking extension is freely available in the 3D Slicer Extension Manager or at https://github.com/koegl/SlicerMRUSLandmarking.

## Code availability

All software used for pre-processing, de-identification, and data conversion of the NRRD images are based on publicly available tools. Specifically, the de-facing algorithm is publicly available at https://github.com/ReubenDo/pydeface-niftyreg and based on the publicly available tools SimpleITK^29^ and NiftyReg^24^. Data conversion was performed using publicly available software tools: 3D Slicer^25^, dicom3tools^26^, and DCMqi^27^. Moreover, we provide scripts to easily convert the dataset from DICOM format into NIfTI or NRRD format, to ensure that our data can be utilized with a wide range of tools. All the scripts for processing and converting the data are available at https://github.com/ReubenDo/ReMIND/.

## Acknowledgements

We would like to acknowledge support from the NIH grants R01EB032387, R01EB027134, P41EB015902, and P41EB028741. We would also like to acknowledge the support and contribution of our collaborating neurosurgeons within the Department of Neurosurgery at Brigham and Women’s Hospital (Boston, USA): Ennio Antonio Chiocca, MD Ph.D.; Timothy R. Smith, MD Ph.D. MPH; and Omar Arnout, MD.

## Author contributions statement

Conceptualization: SF, AJG, TK, WMW Patient recruitment, obtaining consent, performing surgery: AJG, WLB, LR Intraoperative Data Collection: AJG, WLB, PJ, ET, CG, SF; Medical Records Data Collection: CG, PJ Software Design and Development: FK, RD, HC. Data Curation: RD, FK, AK Segmentation: PJ, ET Validation: AJG, WLB, PJ, RD, ET Writing— RD, FK, PJ Review and editing: all authors supervision: TK, SF, AJG; All authors have read and agreed to the published version of the manuscript.

## References

1. Bastos, D. C. D. A. et al. Challenges and opportunities of intraoperative 3D ultrasound with neuronavigation in relation to intraoperative MRI. Front. Oncol. 11, 656519 (2021).

2. Dorward, N. L. et al. Postimaging brain distortion: magnitude, correlates, and impact on neuronavigation. J. Neurosurg. 88, 656–662 (1998).

3. Nabavi, A. et al. Serial intraoperative magnetic resonance imaging of brain shift. Neurosurg. 48, 787–798 (2001).

4. Nimsky, C. et al. Quantification of, visualization of, and compensation for brain shift using intraoperative magnetic resonance imaging. Neurosurg. 47, 1070–1080 (2000).

5. Orringer, D. A., Golby, A. & Jolesz, F. Neuronavigation in the surgical management of brain tumors: current and future trends. Expert. Rev. Med. Devices 9, 491–500 (2012).

6. Menze, B. H. et al. The multimodal brain tumor image segmentation benchmark (BRATS). IEEE Trans. Med. Imaging. 34, 1993–2024 (2014).

7. Dorent, R. et al. Learning joint segmentation of tissues and brain lesions from task-specific hetero-modal domain-shifted datasets. Med. Image Anal. 67, 101862 (2021).

8. Xiao, Y. et al. Evaluation of MRI to ultrasound registration methods for brain shift correction: the CuRIOUS2018 challenge. IEEE Trans. Med. Imaging. 39, 777–786 (2019).

9. Dorent, R. et al. Unified Brain MR-Ultrasound Synthesis Using Multi-modal Hierarchical Representations. In Greenspan, H. et al. (eds.) MICCAI 2023, 448–458 (2023).

10. Luo, J. et al. On the Applicability of Registration Uncertainty. In MICCAI 2019, 410–419 (2019).

11. Mercier, L. et al. Online database of clinical MR and ultrasound images of brain tumors. Med. Phys. 39, 3253–3261 (2012).

12. Rivaz, H., Chen, S. J.-S. & Collins, D. L. Automatic deformable MR-ultrasound registration for image-guided neurosurgery. IEEE Trans. Med. Imaging. 34, 366–380 (2014).

13. Gerard, I. J. et al. Towards a second brain images of tumours for evaluation (BITE2) database. In MICCAI 2016, 16–22 (2016).

14. Xiao, Y., Fortin, M., Unsgård, G., Rivaz, H. & Reinertsen, I. REtroSpective Evaluation of Cerebral Tumors (RESECT): A clinical database of pre-operative MRI and intra-operative ultrasound in low-grade glioma surgeries. Med. Phys. 44, 3875–3882 (2017).

15. Behboodi, B. et al. RESECT-SEG: Open access annotations of intra-operative brain tumor ultrasound images. arXiv preprint at 10.48550/arXiv.2207.07494 (2022).

16. Juvekar, P. et al. The Brain Resection Multimodal Imaging Database (ReMIND). The Cancer Imaging Archive, 10.7937/3RAG-D070 (2023).

17. Blumhofer, A. & Seltz, C. Smooth gray-level based surface interpolation for an isotropic data sets (2011).

18. Ellison, D. W. et al. cIMPACT-NOW update 4: diffuse gliomas characterized by MYB, MYBL1, or FGFR1 alterations or BRAF V600E mutation. Acta Neuropathol. 137, 683–687 (2019).

19. Ellison, D. W. et al. cIMPACT-NOW update 7: advancing the molecular classification of ependymal tumors. Brain Pathol. 30, 863–866 (2020).

20. Louis, D. N. et al. cIMPACT-NOW update 6: new entity and diagnostic principle recommendations of the cIMPACT-Utrecht meeting on future CNS tumor classification and grading. Brain Pathol 30, 844–856 (2020).

21. Wesseling, P. & Capper, D. WHO 2016 Classification of gliomas. Neuropathol. Appl. Neurobiol. 44, 139–150 (2018).

22. Grosu, A.-L. et al. Validation of a method for automatic image fusion (BrainLAB System) of CT data and 11C-methionine-PET data for stereotactic radiotherapy using a LINAC: first clinical experience. Int. J. Radiat. Oncol. Biol. Phys. 56, 1450–1463 (2003).

23. Tokuda, J. et al. OpenIGTLink: an open network protocol for image-guided therapy environment. Int. J. Med. Robot. Comput. Assist. Surg. 5, 423–434 (2009).

24. Modat, M. et al. Global image registration using a symmetric block-matching approach. J. Med. Imaging 1, 024003–024003 (2014).

25. Fedorov, A. et al. 3D Slicer as an image computing platform for the Quantitative Imaging Network. Magn. Reson. Imaging 30, 1323–1341 (2012).

26. Clunie, D. A. Dicom3tools Software. https://www.dclunie.com/dicom3tools.html (2018).

27. Herz, C. et al. DCMQI: an open source library for standardized communication of quantitative image analysis results using DICOM. Cancer Res. 77, e87–e90 (2017).

28. Clark, K. et al. The Cancer Imaging Archive (TCIA): maintaining and operating a public information repository. J. Digit. Imaging 26, 1045–1057 (2013).

29. Yaniv, Z., Lowekamp, B. C., Johnson, H. J. & Beare, R. SimpleITK image-analysis notebooks: a collaborative environment for education and reproducible research. J. Digit. Imaging 31, 290–303 (2018).

